# Predicting the risk of incident hypertension and increase in blood pressure: A systematic review of existing prediction models for adults

**DOI:** 10.1101/2023.04.14.23288537

**Authors:** Pravleen K Bajwa, Natalie A Levine, Leonelo E Bautista

**Affiliations:** Department of Population Health Sciences, School of Medicine and Public Health, University of Wisconsin-Madison, Madison, Wisconsin, USA

## Abstract

**Background and aims:** Models that can predict the risk of developing essential hypertension or increase in blood pressure (BP) can be used to identify high risk individuals. We aimed to summarize and assess prediction models developed in the general adult population using longitudinal data, as well as any external validation of such models.

**Methods:** For this systematic review, we searched the literature on Medline and Embase for studies published between database inception and February 5, 2021. We conducted a narrative synthesis of all models and assessed the risk of bias (ROB) in included studies using PROBAST. We also performed a meta-analysis of all external validation studies validating the Framingham hypertension risk model. We excluded models based on cross-sectional data, or those based on specific patient populations.

**Results:** Our review includes 29 articles which contain 42 prediction models and 11 external validation studies of existing prediction models. Among model development studies, only five models performed both internal and external validation. Among the validation studies, only two existing models were externally validated by researchers other than the ones who developed the model. Most models had low ROB in the predictors and outcomes domains, and half had low ROB in the participants domain. However, all had high ROB in the analysis domain due to inappropriate handling of missing data and/or lack of adequate performance measures, which resulted in high overall ROB for all models.

**Conclusions:** All current risk prediction models predicting hypertension or increased blood pressure have high ROB and most have not been externally validated. New studies should aim to reduce their ROB using standard reporting guidelines and externally validated existing models.

## INTRODUCTION

A systolic blood pressure (SBP) greater than 115 mmHg is the leading risk factor for mortality and morbidity. It is responsible for 9.3% of disability adjusted life years (DALYs) and 10.8 million deaths in the world every year.^1^ Randomized trials have shown that hypertension is preventable even in individuals who already have elevated blood pressure (BP).^2–4^ However, the early identification of individuals at high risk, based on demographic and clinical factors, is key for preventing hypertension.^5^ While a single risk factor can provide a point of reference, combining various risk factors can offer a more accurate risk assessment. A multivariable model can be used to identify high-risk individuals that would benefit from preventive interventions and in designing clinical trials to evaluate new interventions.

Previous systematic reviews of hypertension risk prediction models have summarized their accuracy, commented on the value and practicality of adding genetic risk factors to traditional risk factors, and compared models that were developed using traditional regression methods to those developed using machine learning methods.^6–8^ All previous reviews considered hypertension as a binary outcome but did not address change in BP as a continuous variable. ^6,7^ We expanded on previous reviews by including studies that had prospective change in BP as the model outcome. Additionally, we assessed each study for risk of bias (ROB) using the Prediction model Risk Of Bias Assessment Tool (PROBAST), reported on their adherence to prediction model reporting guidelines, and used a quality assessment framework to ensure that our review is transparent and reproducible.^9–11^ Overall, our objective was to identify, summarize, and assess the ROB in published risk prediction models for incident hypertension or increase in BP, as well as any studies validating such models.

## METHODS

We used the Preferred Reporting for Systematic Reviews and Meta-Analyses (PRISMA) for our review.^12^ The protocol for this systematic review was registered on PROSPERO (CRD42021224240) in January 2021 and can be accessed at www.crd.york.ac.uk/PROSPERO/display_record.asp?ID=CRD42021224240.13

### Eligibility criteria

#### Population

Studies on adults (age ≥ 18 years) were included. Studies on specific patient populations, such as hospitalized and pregnant individuals, who could have secondary instead of primary hypertension, were not eligible.

#### Outcomes

We included all studies reporting the development or validation of a risk assessment model, equation, score, or tool for prediction of essential hypertension, prehypertension, or BP change. We excluded studies with only one predictor, studies with only current BP measures as predictors, risk association studies, and those predicting secondary hypertension (hypertension that presents with a comorbidity associated with an increased risk of hypertension risk, e.g., diabetes, chronic kidney disease) and gestational hypertension.

#### Study design

We were interested in models that predict future BP outcomes using baseline data. Therefore, we only considered studies based on longitudinal data.

#### Publication type

All English language studies published by the date of our search (February 5, 2021) were included. There were no limits set on the length of follow-up in a study.

#### Search strategy

We used Medline (via Pubmed) and Embase for our systematic review, with the final search on February 5, 2021. We developed an initial search strategy based on known relevant studies and our subject matter knowledge. We then added additional terms using Pubmed’s MeSH database. Our final strategy was reviewed by our institution’s Health Sciences librarian and validated by testing whether it could identify seven known relevant studies. A comprehensive search strategy for both databases is available in Supplement 1.

### Study selection

Two reviewers (PB, NL) independently screened retrieved articles for eligibility based first on title, then on the abstract for those not excluded thus far, and finally based on the full text for those not excluded during the abstract screening. Disagreements were assessed after the full-text screening and resolved by a third reviewer (LB) when needed.

### Data extraction

Two reviewers (PB, NL) extracted items from the selected studies using a pilot-tested form. The list of items was based on the Cochrane guidance for data extraction and the CHecklist for critical Appraisal and data extraction for systematic Reviews of prediction modeling Studies (CHARMS) and included participant characteristics, study design, predictors considered, statistical model used, model performance, and type of validation performed.^11,14^ We also evaluated whether articles published after 2015 adhered to the Transparent Reporting of a multivariable prediction model for Individual Prognosis Or Diagnosis (TRIPOD) guidelines.^10,15^ The full list of extracted items is included in Supplement 2. If an article included multiple models, we conducted separate data extraction for each model. We also extracted summary measures including sample size, total follow-up, number of events, event horizon, discrimination measures, and calibration measures (Box 1).

##### Box 1: Definitions of technical terms

**Internal validation**: Reusing parts or all of the dataset on which a model was developed to assess overfitting and correct for the resulting ‘optimism’ in the performance of the model, e.g., cross validation, bootstrapping.^46^

**External validation**: Assessing whether the model predictions hold true in a setting different than the one of its development sample.^44^

**Discrimination**: The ability of a prediction model to differentiate between those who do or do not experience the outcome event.^47^

**Calibration**: Agreement between the predicted outcomes from the model and the observed outcomes.^47^

### Risk of bias in individual studies

Models with limitations in study design, conduct, or analysis can have biased findings and result in poorer predictive performance (internal validity). To evaluate this risk, we used PROBAST, which has been designed to assess ROB in prediction model studies in 4 domains – participants, predictors, outcome, and analysis.^9^

### Synthesis of results

We conducted a narrative synthesis of data extracted from the studies and have presented it in tabular form. For studies that had more than three models, we selected the best models (as indicated by the study authors) or the ones with the most information. Due to the heterogeneity in the number and types of predictors, methods, study design, and settings of the included studies, an informative meta-analysis was not feasible. The Framingham Hypertension Risk model (FHR) was the only model that has been validated in various populations.^16^ The studies validating the FHR model were directly comparable and illustrated its applicability in different populations. Therefore, we conducted a meta-analysis with the AUC estimates from each model that validated the FHR model (Supplement 3). All analyses were conducted in Stata 17 (StataCorp LLC, College station, TX).

## RESULTS

Our literature search resulted in 14,328 records (Figure 1). After removing duplicates, we screened 9,710 records and assessed 69 full-text articles for eligibility. After excluding 40 articles that did not meet our inclusion criteria, 29 articles were eligible for the review.

**Figure 1.**
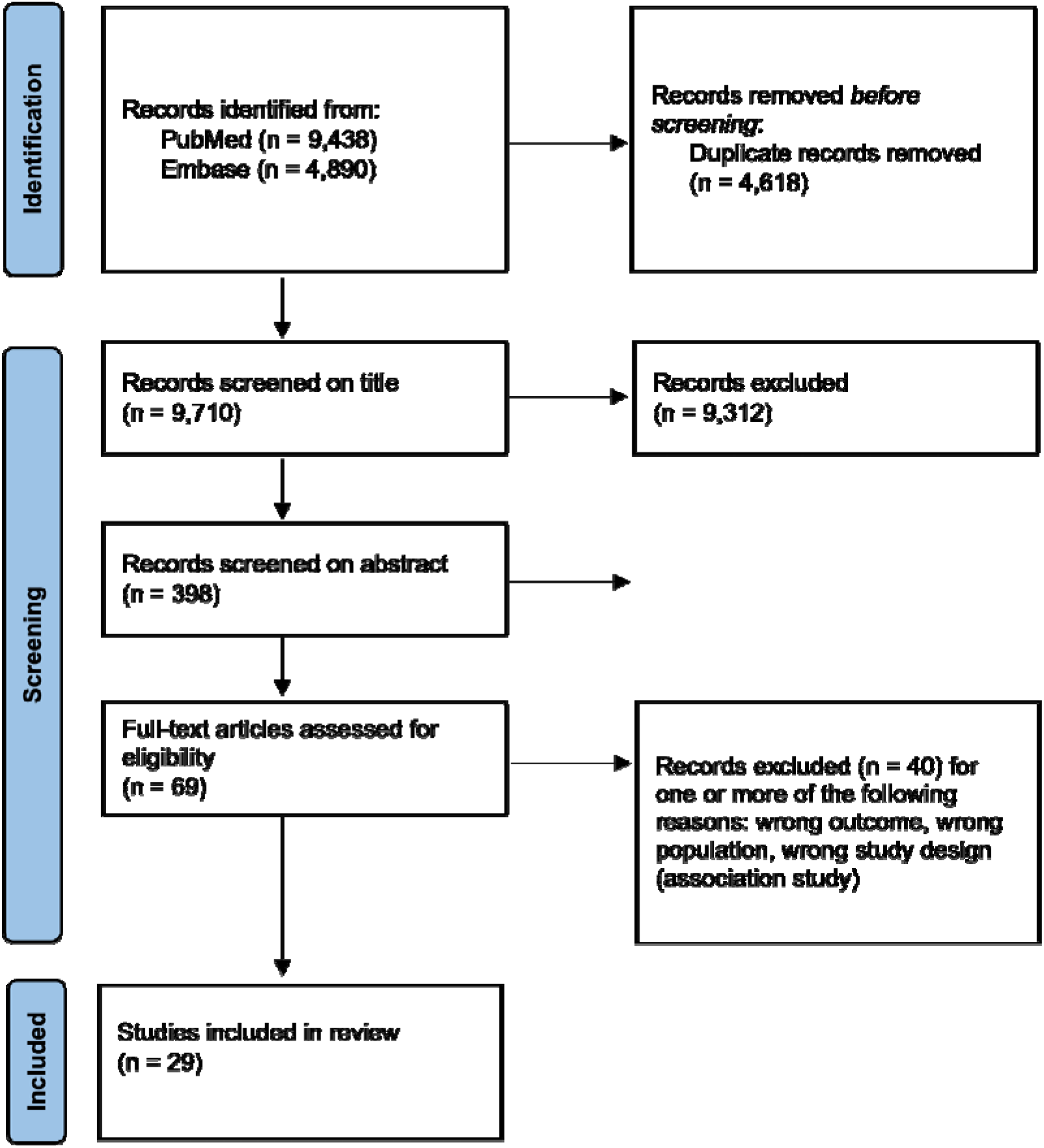
Study selection for the systematic review (PRISMA^12^ flow diagram)

### Study characteristics

#### Model development studies

Out of the 29 articles included in our review, 25 described developing a new prediction model. These 25 studies developed 42 new models for predicting the risk of hypertension or increase in BP (Table 1). Sixteen studies developed models using data from Asian countries, five from European countries, and four from North American countries. The number of participants ranged from 297 to 823,627, and their ages ranged from 18 years old to 88 years and older. The duration of follow-up ranged from 1 to 35 years. Two studies had only male participants,^17,18^ one study had only female participants,^19^ and the rest had both sexes.

**Table 1.** Characteristics of studies included in the review

| First author, year | Model or study name(s), if given | Country | Population | % Women | Age range | Race/ ethnicity | Length of follow-up | Outcome | Outcome definition | Sample size | Cases /N* |
| --- | --- | --- | --- | --- | --- | --- | --- | --- | --- | --- | --- |
| Pearson, 1990 | Johns Hopkins Multiple Risk Equations | USA | Johns Hopkins medical students in 1984 who participated in the Preventive Cardiology Academic Award Collaborative Project | 0 | 22-26 | White | 19-35 years | Incident hypertension | Elevated BP requiring pharmacologic therapy (self-reported) | 1,130 | 10.1 |
| Parikh, 2008 | Framingham Risk Score-Near Term Hypertension | USA | Offspring of original Framingham cohort | 54.1 | 20-69 | White | 3.8 years median | Incident hypertension | SBP ≥ 140 mmHg or DBP ≥ 90 mmHg; BP-lowering medication | 1,717 | 46.4 |
| Kivimaki, 2009 | Whitehall II Risk Score | England | Civil servants in London | 31 | 35-55 | Mostly White (92%) | 5.6 years | Incident hypertension | SBP ≥ 140 mmHg or DBP ≥ 90 mmHg; antihypertensive medication | 6,704 | 30.5 |
| Paynter, 2009 | Women's Health Study | USA | Female health professionals followed prospectively for incident cardiovascular disease, including hypertension | 100 | 45+ | Mostly white (96%) | 8 years | Incident hypertension | Physician diagnosis at year 1, 3, or annually thereafter; New hypertensive treatment at year 1, 3, or 4; SBP ≥ 140 mmHg at year 1 or 4; DBP ≥ 90 mmHg at year 1 or 4 | 14,822 | 20.3 |
| Kivimaki, 2010 | Whitehall II Repeat-measure Risk Score, Average BP Risk Score, Usual BP Risk Prediction Score | England | Civil servants in London | 30.1 | 35-55 | Mostly white | 5.8 years median | Incident hypertension | SBP ≥ 140 mmHg or DBP ≥ 90 mmHg; antihypertensive medication | 4,314 | 24.4 |
| Bozorgmanesh, 2011 | Tehran Lipid and Glucose Study | Iran | Residents of Tehran | 57.9 | 20+ | Iranian | 6 years | Incident hypertension | SBP ≥ 140 mmHg or DBP ≥ 90 mmHg; self- reported BP-lowering medication | 4,656 | 17.3 |
| Chien, 2011 | - | Taiwan | Residents of one township | 50.8 | 35+ | Chinese | 6.15 years median | Incident hypertension | SBP ≥ 140 mmHg or DBP ≥ 90 mmHg; anti-hypertensive medications | 2,506 | 41.1 |
| Tseng, 2011 | Keelung Community-based Integrated Screening (KCIS) | Taiwan | Residents of Keelung | 63.2 | Adults | Majority Chinese | 3 years | Progression or regression of hypertension | Normotension: SBP < 120 mmHg and DBP < 80 mmHg; Prehypertension: SBP 120-139 mmHg or DBP 80-89 mmHg; Stage 1 hypertension: SBP 140-159 mmHg or DBP 90-99 mmHg; Stage 2 hypertension: SBP ≥ 160 mmHg or DBP ≥ 100 mmHg | 42,027 | NR |
| Fava, 2013 | Swedish genetic risk score (BP-GRS29); Malmo Preventive Project (MPP) | Sweden | Urban Swedish residents of Malmo | 36.7 | 45.2± 7.4 | Swedish | 23 years mean | Incident hypertension, change in BP | SBP ≥ 140 mmHg or DBP ≥ 90 mmHg; antihypertensive treatment | 18,240 | 63.3 |
| Lim, 2013 | Korean Genome and Epidemiology Study (KoGES) | South Korea | Residents of Ansan (industrialized) and Ansong (rural) regions | 52.1 | 40-69 | Korean | 4 years | Incident hypertension | SBP ≥ 140 mmHg or DBP ≥ 90 mmHg; antihypertensive medication | 4,747 | 17.3 |
| Volzke, 2013 | Study of Health in Pomerania (SHIP) | Germany | Sample was drawn from population registries | 63.1 | 20-79 | White | 5 years | Incident hypertension | SBP ≥ 140 mmHg or DBP ≥ 90 mmHg; antihypertensive medication | 1,605 | 20.1 |
| Lim, 2015 | Korean Genome and Epidemiology Study (KoGES) | South Korea | Residents of Ansan (industrialized) and Ansong (rural) regions | 51.5 | 40-69 | Korean | 4 years | Incident hypertension | SBP ≥ 140 mmHg or DBP ≥ 90 mmHg; antihypertensive medication | 5,632 | NR |
| Otsuka, 2015 | - | Japan | Employees at an electrical equipment manufacturing company in | 0 | 19-63 | Japanese | 4 years median | Incident hypertension | SBP ≥ 140 mmHg or DBP ≥ 90 mmHg; antihypertensive medication | 15,025 | 10.9 |
|  |  |  | Japan |  |  |  |  |  |  |  |  |
| Zhang, 2015 | Hypertension Synthetic Predictor (HSP) | China | Those who received annual medical examinations from 2005 through 2010 at 2 hospitals | 41.4 | 18-88 | Han Chinese | 5 years | Incident hypertension | SBP ≥ 140 mmHg or DBP ≥ 90 mmHg; anti-hypertensive medication | 17,471 | 21.7 |
| Chen, 2016 | - | China | Those who participated in routine health check-ups | 39.7 | 18+ | Han Chinese | 4 years median | Incident hypertension | SBP ≥ 140 mmHg or DBP ≥ 90 mmHg; antihypertensive medication | 12,497 | 22.3 |
| Lee, 2016 | National Health Insurance Corporation (NHIC) cohort in South Korea | South Korea | Those who went to health check-ups | 46 | 20-69 | Korean | 2 years | Incident hypertension | Medical treatment record with the diagnosis code for hypertension; SBP ≥ 90mmHg or DBP ≥ 140 mmHg | 148,890 | 7.0 |
| Ramezankhani, 2016 | Tehran Lipid and Glucose Study | Iran | Residents of district 13 in Tehran, under the coverage of 3 medical centers | 55.5 | 20+ | Iranian | 8.7 years median | Incident hypertension | SBP ≥ 140 mmHg or DBP ≥ 90 mmHg; antihypertensive medications | 6,205 | 23.6 |
| Sathish, 2016 | - | India | Rural Indians | 52.2 | 15-64 | Indian | 7.1 years | Incident hypertension | SBP ≥ 140 mmHg or DBP ≥ 90 mmHg; BP-lowering medications | 297 | 23.6 |
| Kanegae, 2018 | The Jichi Genki hypertension prediction model (JG model) | Japan | Members of organizations and employees of companies in Tokyo | 47.8 | 18-83 | Japanese | 3.4 years mean | Incident hypertension | SBP ≥ 140 mmHg or DBP ≥ 90 mmHg; initiation of antihypertensive medication with self-reported hypertension | 63,495 | 11.7 |
| Wang, 2018 | China Health and Nutrition Survey | China | General population | 63.0 | 18-90 | Chinese | 8.05±5.27 years mean | Transition between normotensive, pre-hypertensive, hypertensive states | Normotension: No antihypertensive drugs, SBP < 120 mmHg and DBP < 80 mmHg; Pre-hypertension: No antihypertensive drugs, SBP 120–139mmHg (DBP < 90mmHg) or DBP 80–89mmHg (SBP | 5,265 | 16.8* |
| | | | | | | | | | < 140mmHg);<br>Hypertension:<br>Antihypertensive drugs<br>or SBP ≥ 140mmHg or<br>DBP ≥ 90 mmHg.<br>Censored once<br>hypertensive | | |
| Ye, 2018 | - | USA | Patients from 35 hospitals, 34 federally qualified health centers, 400+ ambulatory practices in the state of Maine from 2013 to 2015 | 55.5 | NR | Mostly white (95%) | 1 year | Incident hypertension | ICD-9-CM diagnosis codes from category 401 in a patient's medical record | 823,627 | 11.6 |
| Kanegae, 2019 | - | Japan | Those who underwent health checkups | 55.4 | 46.4± 12.1 | Japanese | 2 years | Incident hypertension | SBP ≥ 140 mmHg or DBP ≥ 90 mmHg; initiation of antihypertensive medication with self-reported hypertension | 18,258 | 14.6 |
| Xu, 2019 | - | China | Permanent residents of rural areas of Henan Province | NR | 35-74 | Chinese | 6 years | Incident hypertension | SBP ≥ 140 mmHg or DBP ≥ 90 mmHg; diagnosis by a physician and current anti-hypertension treatment | 8,319 | 12.5 |
| Giontella, 2021 | BP-GRS858; Malmo Preventive Project (MPP) | Sweden | Urban Swedish residents of Malmo | NR | NR | Swedish | 23 years mean | Incident hypertension | SBP ≥ 140 mmHg or DBP ≥ 90 mmHg; antihypertensive drugs | 6,021 | NR |
| Wang, 2021 | The Rural Chinese Cohort Study | China | Adults from a rural district using cluster random sampling | 60.7 | 18-70 | Chinese | 6 years | Incident hypertension | SBP ≥ 140 mmHg or DBP ≥ 90 mmHg; BP-lowering medications | 9,034 | 18.4 |

\*Cases = Incident hypertension. Note that some studies were looking at transition between BP states, not just incident hypertension. All states are not captured in this column.

Most models used traditional regression methods like Cox proportional hazards (n=11), Weibull regression (n=10), and logistic regression (n=10) (Table 2). Seven models used machine learning methods,^20–23^ two used Bayesian Network Analysis,^24,25^ and one model used a multistate Markov model.^26^ Summary information for each model in the 25 studies is presented in Table 2. As some studies had multiple models, we will use the model numbers listed in Table 2 to reference models throughout the paper.

**Table 2.**
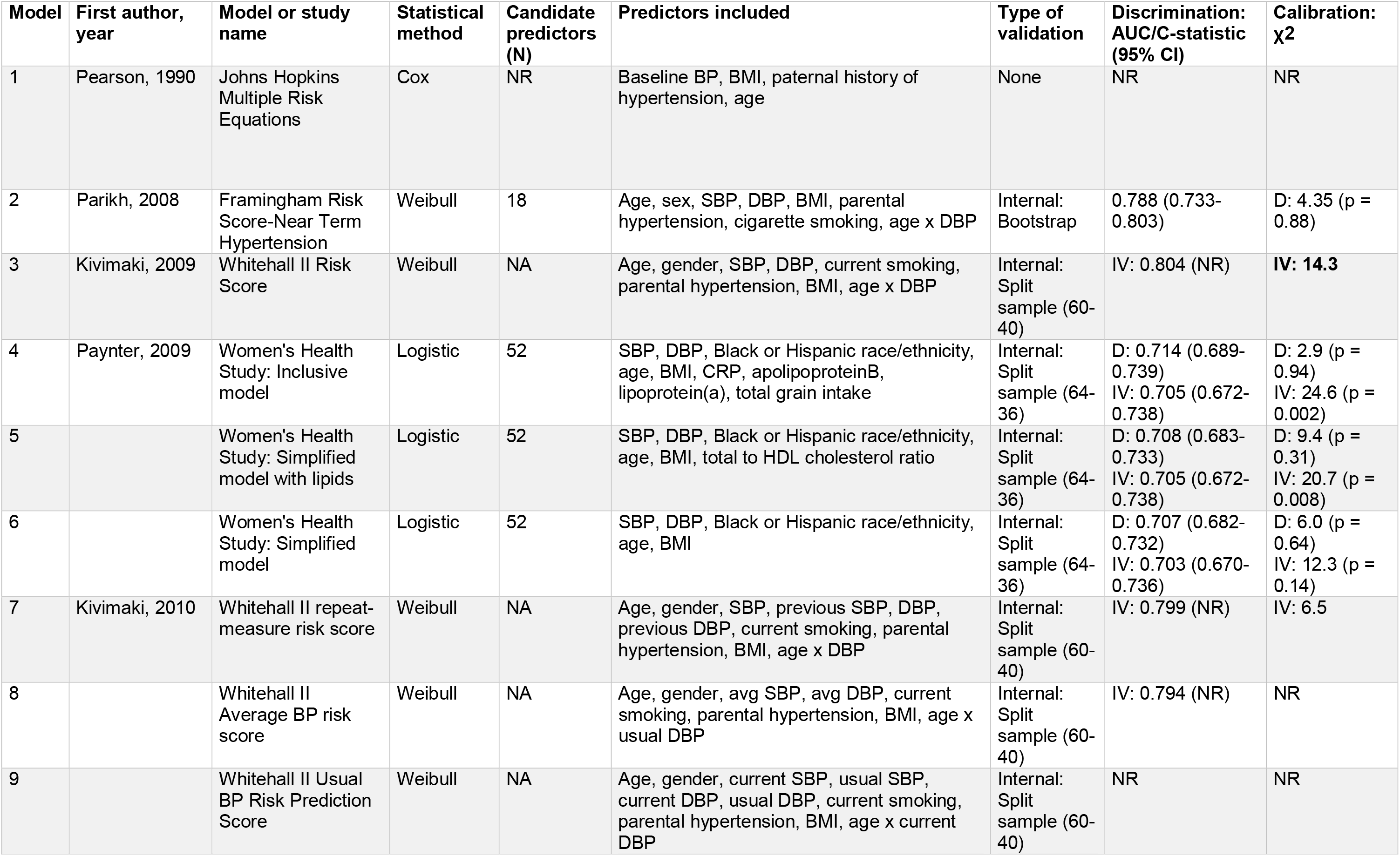
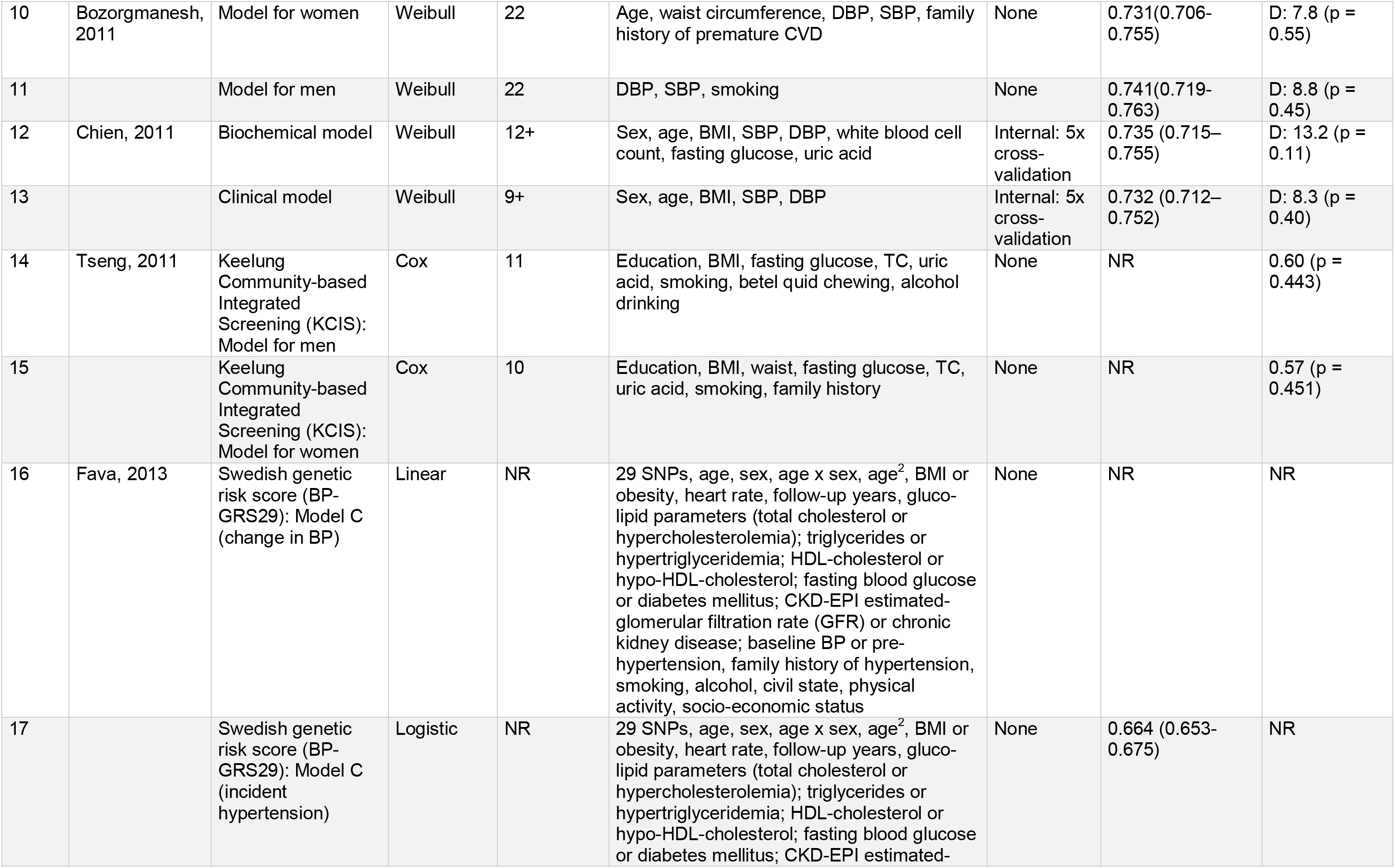
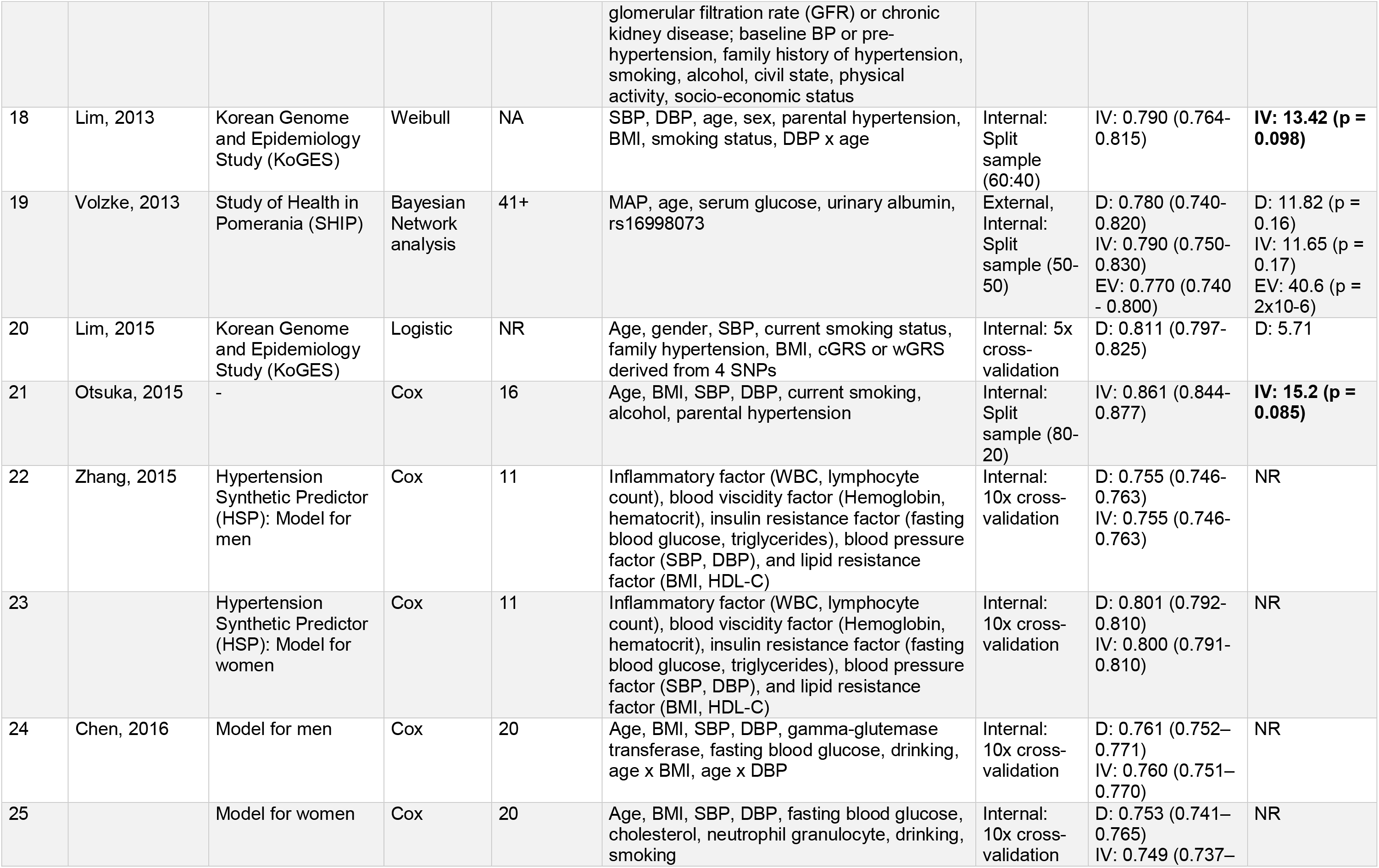
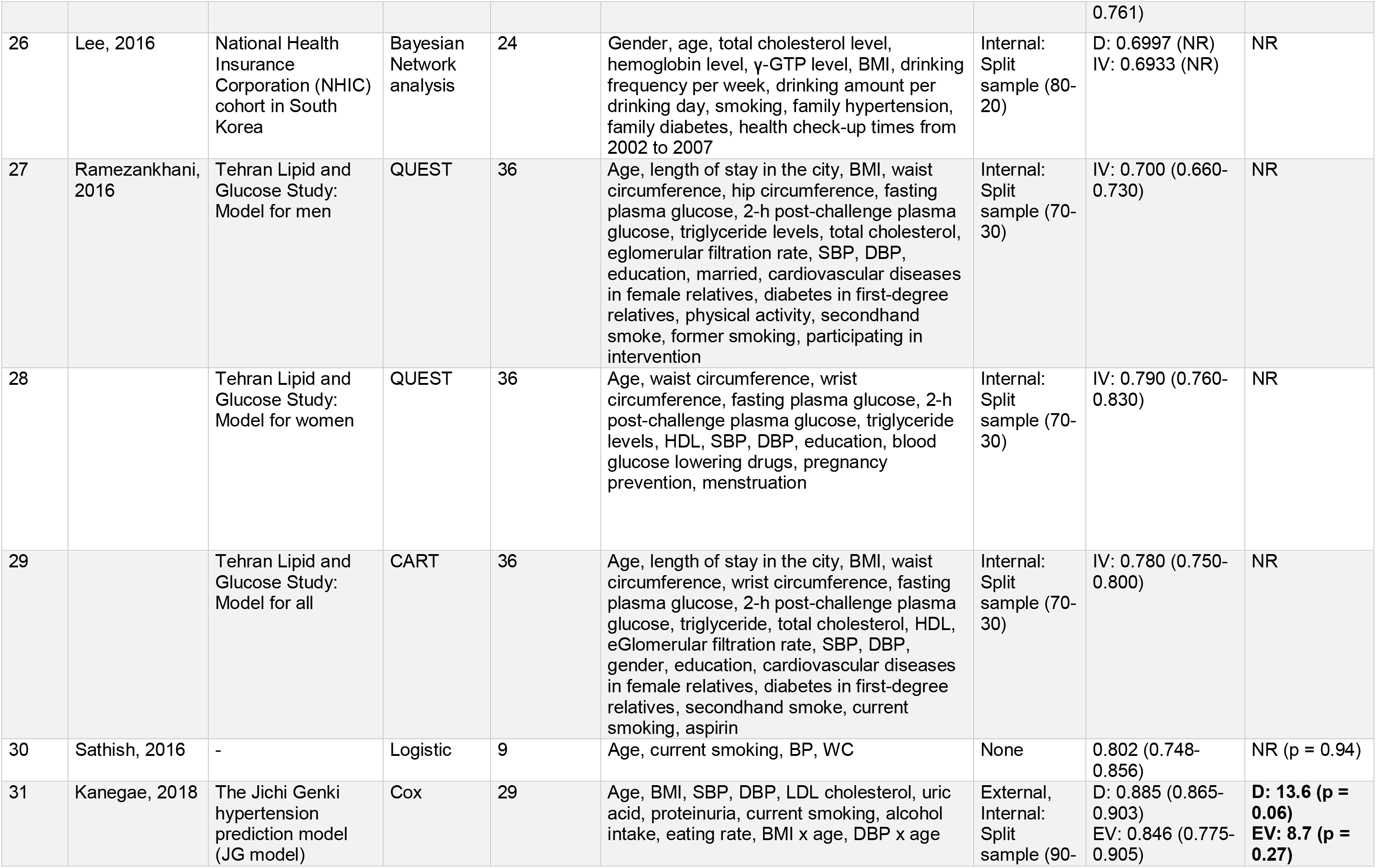
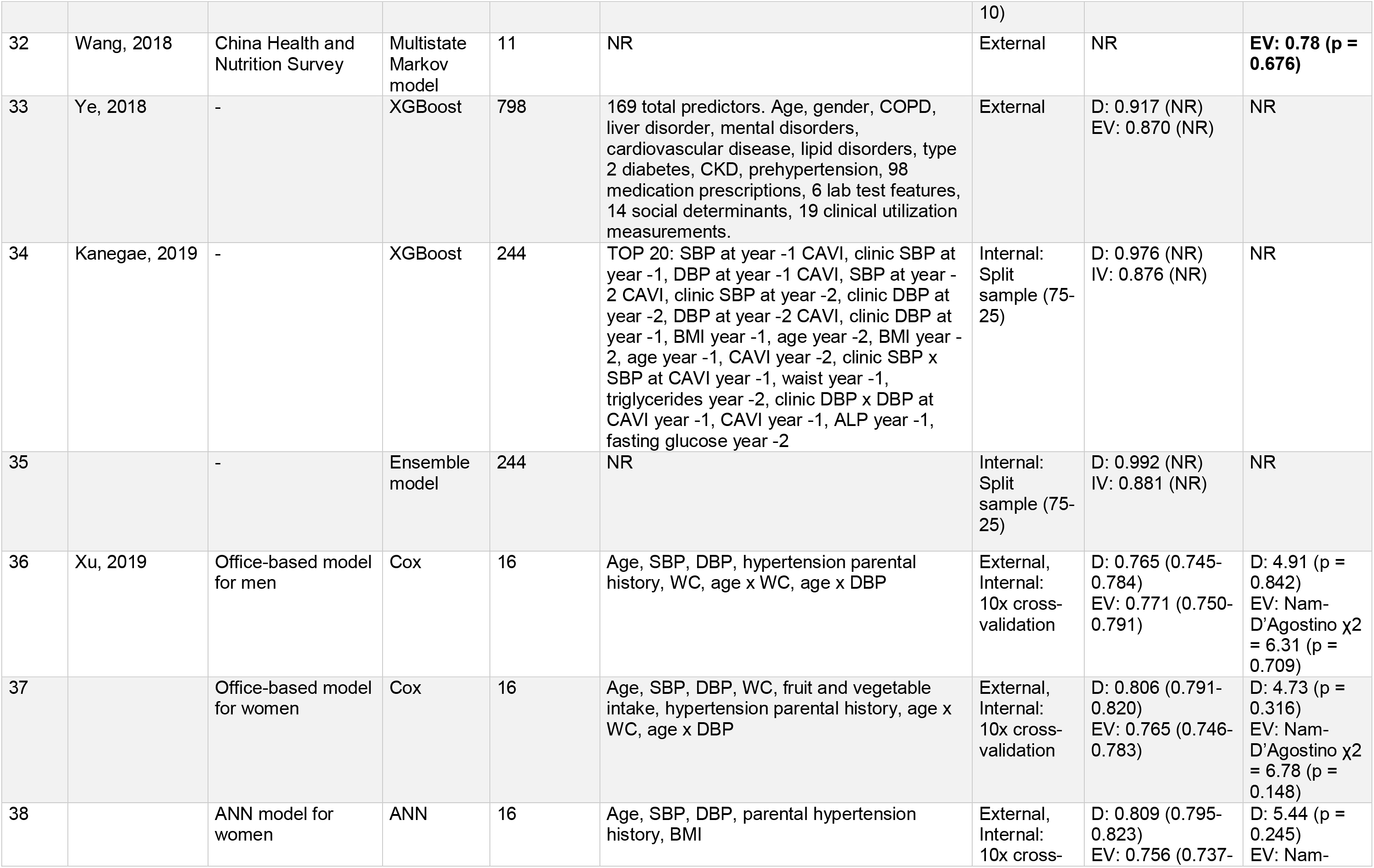
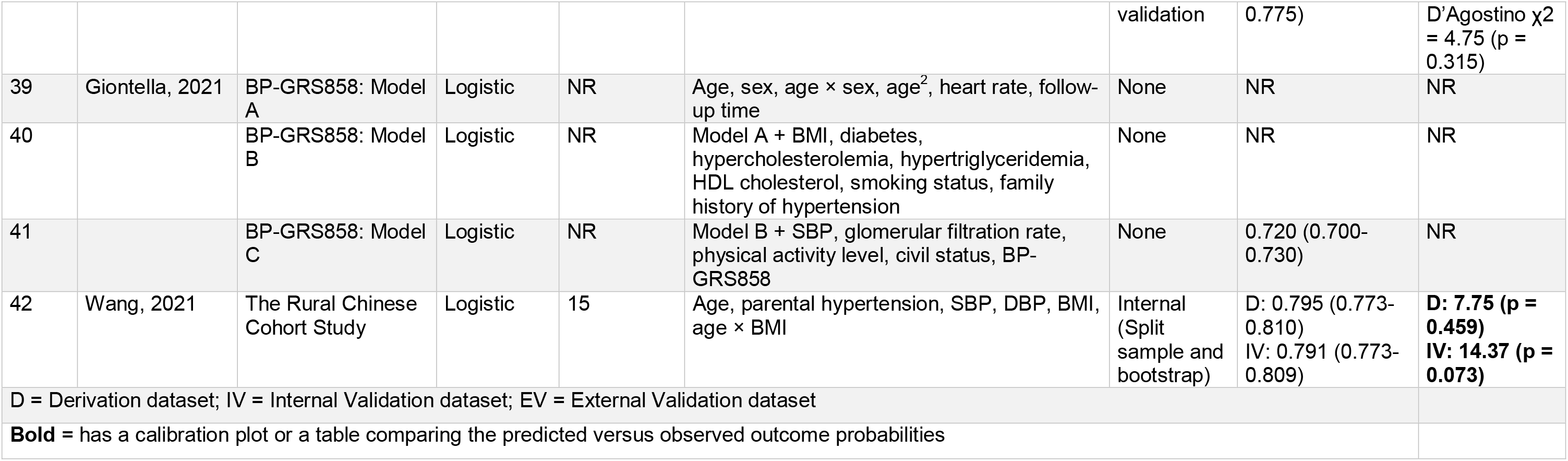
Development of models predicting hypertension or increased blood pressure

Most models were developed with incident hypertension as the outcome. One study included models for change in SBP and DBP, in addition to incident hypertension (Table 2: Model 16).^27^ One study modeled the progression or regression of hypertension as the outcome (Table 2: Models 14-15), and another modeled the transition between normotensive, pre-hypertensive, and hypertensive status (Table 2: Model 32).^28,29^ All studies excluded participants with hypertension at baseline. Most studies defined hypertension as SBP ≥ 140mmHg or DBP ≥ 90 mmHg, or the use of antihypertensive medication or treatment. However, Pearson et al.^17^ defined hypertension as self-reported elevated BP requiring medication, Tseng et al.^27^ did not include taking antihypertensive medication in their definition, and ^21^et al. used ICD-9-CM codes acquired from electronic health records.

Most (n = 36) models shared information about candidate variables considered for their prediction model (range: 9 to 798). All except two models (Table 2: Models 32 & 35) shared information about the final predictors included in their model.^22,26^ Age, gender, SBP, diastolic blood pressure (DBP), and body mass index were the most frequently included predictors in final models. While only 17 models had gender in their final model, 16 additional models were gender-specific. Furthermore, smoking, and a family history of hypertension were included in close to half the models. Sets of variables that were included in models from at least three studies are presented in Table 3.

**Table 3.** Predictors included in at least three model development papers

| Model | Age | Gender | Education | SBP | DBP | BMI | Waist circumference | Lipids | Glucose | Family history of hypertension | Smoking | Alcohol | Genetic risk score / SNPs |
| --- | --- | --- | --- | --- | --- | --- | --- | --- | --- | --- | --- | --- | --- |
| 1 | - |  |  | - | - | - |  |  |  | - |  |  |  |
| 2 | - | - |  | - | - | - |  |  |  | - | - |  |  |
| 3 | - | - |  | - | - |  |  |  |  | - | - |  |  |
| 4 | - |  |  | - | - | - |  | - |  |  |  |  |  |
| 5 | - |  |  | - | - | - |  | - |  |  |  |  |  |
| 6 | - |  |  | - | - | - |  |  |  |  |  |  |  |
| 7 | - | - |  | - | - | - |  |  |  | - | - |  |  |
| 8 | - | - |  | - | - | - |  |  |  | - | - |  |  |
| 9 | - | - |  | - | - | - |  |  |  | - | - |  |  |
| 10 | - |  |  | - | - |  | - |  |  |  |  |  |  |
| 11 |  |  |  | - | - |  |  |  |  |  | - |  |  |
| 12 | - | - |  | - | - | - |  |  | - |  |  |  |  |
| 13 | - | - |  | - | - | - |  |  |  |  |  |  |  |
| 14 |  |  | - |  |  | - |  | - | - |  | - | - |  |
| 15 |  |  | - |  |  | - |  | - | - | - | - |  |  |
| 16 | - | - |  | - | - | - |  | - | - | - | - | - | - |
| 17 | - | - |  | - | - | - |  | - | - | - | - | - | - |
| 18 | - | - |  | - | - | - |  |  |  | - | - |  |  |
| 19 | - |  |  |  |  |  |  |  | - |  |  |  | - |
| 20 | - | - |  | - |  | - |  |  |  | - | - |  | - |
| 21 | - |  |  | - | - | - |  |  |  | - |  |  |  |
| 22 |  |  |  | - | - | - |  | - | - |  |  |  |  |
| 23 |  |  |  | - | - | - |  | - | - |  |  |  |  |
| 24 | - |  |  | - | - | - |  |  | - |  | - | - |  |
| 25 | - |  |  | - | - | - |  | - | - |  | - | - |  |
| 26 | - | - |  |  |  | - |  |  |  | - | - | - |  |
| 27 | - |  | - | - | - | - | - | - | - |  |  |  |  |
| 28 | - |  | - | - | - | - | - | - | - |  |  |  |  |
| 29 | - | - | - | - | - | - | - | - | - |  | - |  |  |
| 30 | - |  |  | - | - |  | - |  |  |  | - |  |  |
| 31 | - |  |  | - | - | - |  | - | - |  | - | - |  |
| 32 |  |  |  |  |  |  |  |  |  |  |  |  |  |
| 33 | - | - | - |  |  |  |  |  |  |  |  |  |  |
| 34 | - |  |  | - | - | - |  | - | - |  |  |  |  |
| 35 |  |  |  |  |  |  |  |  |  |  |  |  |  |
| 36 | - |  |  | - | - |  | - |  |  | - |  |  |  |
| 37 | - |  |  | - | - |  | - |  |  | - |  |  |  |
| 38 | - |  |  | - | - | - |  |  |  | - |  |  |  |
| 39 | - | - |  |  |  |  |  |  |  |  |  |  |  |
| 40 | - | - |  |  |  | - |  |  |  | - | - |  |  |
| 41 | - | - |  | - |  | - |  | - |  | - | - |  | - |
| 42 | - |  |  | - | - | - |  | - |  | - |  |  |  |
| **Total** | **35** | **17** | **6** | **33** | **31** | **31** | **7** | **16** | **15** | **19** | **20** | **7** | **5** |

#### Model validation studies

Out of the 29 articles included in our review, nine externally validated an existing hypertension prediction model (Table 4).^24,30–37^ The number of participants ranged from 802 to 24,434, between 18 and 84 years of age, and were followed for 1.6 years to 25 years. All nine articles validated the FHR model and one additionally validated the Johns Hopkins multiple risk equations as well.^32^ The FHR model was originally developed using data from primarily White participants recruited in Framingham, Massachusetts (USA).^16^ It has been externally validated in populations from the United Kingdom,^30^ Taiwan,^32^ South Korea,^34^ Germany,^24^ Denmark,^24^ China,^35,37^ and Brazil,^36^ and more ethnically diverse cohorts in the USA.^31,33^ The Hopkins model,^17^ which was also originally developed using data from the USA, was validated in a population from Taiwan.^32^

**Table 4.** External validation of existing* hypertension prediction models.

| **First author, year** | **Model validated** | **Validation population** | **Race or ethnicity** | **Age range** | **Length of follow-up** | **Sample size** | **Cases** | **Discrimination: AUC/C-statistic (95% CI)** | **Calibration: HL $\chi^2$ (p-value)** |
| --- | --- | --- | --- | --- | --- | --- | --- | --- | --- |
| Kivimaki, 2009 | Framingham | UK; London office workers | Mostly (92%) White | 35-55 | 5.6 years median | 5472 person-examinations | 785 | 0.803 (NR) | **11.5 (NR)** |
| Muntner, 2010 | Framingham | USA; MESA | Caucasian, African-American, Hispanic, and Asian - primarily of Chinese descent | 45-84 | 1.6 years median | 3013 | 849 | 0.788 (0.773-0.804) | **NR (p < 0.001)** |
| Chien, 2011 | Framingham | Taiwan; one township | Chinese | 35 and older | 6.15 years median | 2506 | 1029 | 0.709 (0.688-0.730) | 7.4 (p = 0.49) |
| Chien, 2011 | Johns Hopkins | Taiwan; one township | Chinese | 35 and older | 6.15 years median | 2506 | 1029 | 0.707 (0.686-0.728) | 16.7 (p = 0.03) |
| Carson, 2013 | Framingham | USA; 4 urban cities | White, Black | 18-30 | 25 years | 4388 | 1179 | 0.84 (0.83-0.85) | After recalibration using CARDIA = 14.6 (p = 0.067)<br>Before recalibration = 249.4 (p < 0.001) |
| Lim, 2013 | Framingham | Korea; KoGES | Korean | 40-69 | 4 years | 1907 | 336 | 0.789 (0.764-0.815) | **29.73 (p = 0.0002)** |
| Volzke, 2013 | Framingham | Germany; SHIP | White | 20-79 | 5 years | 802 | 157 | 0.77 (0.73-0.82) | 11.26 (p = 0.19) |
| Volzke, 2013 | Framingham | Denmark; INTER99 | White | 30-60 | 5 years | 2887 | 434 | 0.73 (0.71-0.75) | 203.34 (p < 10<sup>-8</sup>) |
| Zheng, 2014 | Framingham | China; Rural areas of one county | Chinese | 35 and older | 4.8 years median | 24434 (105,702 person-years) | 8675 | 2 year: 0.537 (0.524-0.550)<br>4 year: 0.610 (0.602-0.618) | **2-year = 2,287.7 (p < 0.0001)**<br>**4-year = 8,227.1 (p < 0.0001)** |
| Sylos, 2020 | Framingham | Brazil; ELSA-Brasil | Brazilian - primarily White, Brown, and Black | 35-70 | 4 years | 3202 | NR | 0.827 (0.808-0.847) | 3.78; (p = 0.876) |
| Wang, 2021 | Framingham | China; Rural districts | Chinese | 18-70 | 6 years median | 3551 | NR | 0.787 (0.778-0.795) | NR |

### Model performance

Most models (n = 34; 81.0%) reported discrimination measures (AUC), ranging from 0.66 (95% CI: 0.65-0.82) to 0.99 (95% CI not reported). For calibration, 23 (54.8%) models reported a χ2 statistic, 6 (14.3%) additionally presented a calibration curve or another visual representation, and 19 (45.2%) did not report calibration at all (Table 2). Out of 42 models, 24 performed only internal validation, 2 performed only external validation, 5 performed both, and 11 performed none.

Among model validation papers, all models except one^37^ have calibration measures or a visual representation of calibration. In the random-effects meta-analysis of models that validated the FHR model, we found that the pooled AUC was 0.758 (95% CI: 0.692-0.825) overall, 0.783 (95% CI: 0.736-0.830) in European and North American populations, 0.735 (95% CI: 0.604-0.866) in Asian and South American populations (Supplement 3). Significant heterogeneity was present even when categorized geographically (I^2^ = 99.4% overall, 96.4% Europe and North America, 99.7% Asia and South America).

### Quality Assessment

Most models in the model development studies had low ROB in the domains of predictors and outcomes (71.4%). In the participants domain, 60.0% of the models had low ROB, and 23.8% had high ROB due to use of an inappropriate data source (n = 6) or inappropriate inclusion and exclusion criteria (n = 4). All models showed high ROB in the analysis domain mainly due to incorrect handling of observations with missing data (n = 35), not including all enrolled participants in the analysis (n = 39), and not evaluating model performance measures (n = 37). All models were classified as having high overall ROB due to one or more of the domains having high ROB (Figure 2A).

**Figure 2.**
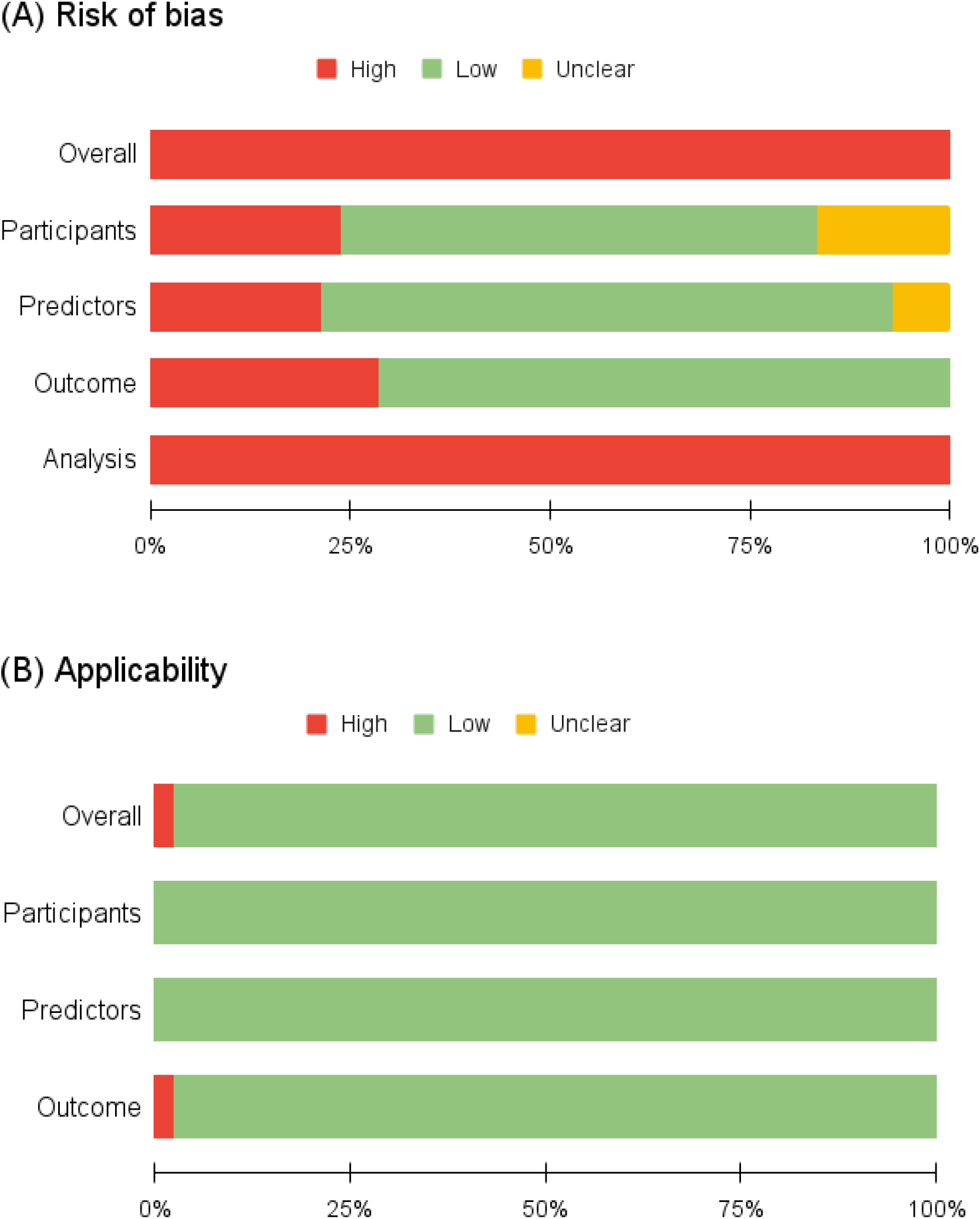
Risk of bias (A) and applicability (B) in hypertension prediction models.

For the applicability assessment, 41 models were considered to have low applicability concerns based on the participants, predictors, and outcome domains (Figure 2B). One model had high overall risk of applicability due to concerns with outcome definition (Table 2: Model 1).^17^ Among model development studies published after 2015, only 1 out of 19 models met TRIPOD guidelines (Table 2: Model 31).^38^

All model validation studies had low ROB based on the participants, predictors, and outcome domains. All models had high ROB in the analysis domain, due to inappropriate handling of observations with missing data (n = 11) and not including all enrolled participants in the analysis (n = 11). Full details of each model’s quality assessment are available in Supplement 4. Among model validation studies published after 2015, neither of models met TRIPOD guidelines.

## DISCUSSION

This systematic review summarizes 29 published studies on risk prediction models for hypertension or increase in BP, which includes 42 models as well as nine external validations of existing models. It also reports on their ROB according to the PROBAST tool.

### Explanation of major findings

Most models were developed using East Asian, European, and North American populations. None were developed using data from Africa, South America, Central America, or Oceania. There was no difference in the types of predictors used in models from distinct populations. Most models used predictors that are readily available in a healthcare or research setting, making the models practical. Some also incorporated genetic variants, but we found this resulted in only a modest increase in discrimination. For example, an increase in AUC from 0.662 (95% CI: 0.651-0.672) to 0.664 (0.653-0.675) in Fava et al. 2013, and from 0.810 (95% CI: 0.796-0.824) to 0.811 (95% CI: 0.797-0.825) in Lim et al. 2015 was observed after adding genetic components to the model.^28,39^ This modest improvement in model discrimination is consistent with findings from a recent systematic review of models predicting cardiovascular risk.^40^

Only one model predicted change in BP (Table 2: Model 16)^28^ and the rest defined hypertension by dichotomizing BP cut-offs or using self-reported physician diagnosis. The latter could limit the usefulness of the prediction models in the future as the cut-points used to define hypertension have changed and may continue to do so in the future. All models that reported their BP cut-off values defined SBP ≥ 140 mmHg or DBP ≥ 90 mmHg as hypertensive. While hypertension thresholds in the ESC/ESH guidelines have not changed over the years, the thresholds used in the US (previously JNC, now ACC/AHA) have.^41^ The latest ACC/AHA guidelines define hypertension as SBP ≥ 130 mmHg or DBP ≥ 80 mmHg, which is distinct from the European thresholds and the thresholds used in the models included in this review.^42,43^

Most authors performed internal validation (n = 29) of their models but only seven conducted an external validation. Twelve models did not undergo any sort of validation. Among those that internally validated their model, split-sample and cross-validation were the most common methods, with only two models using bootstrap.^16,37^ Model performance measures were either based on the derivation dataset (apparent performance) which is over-optimistic, or on internal validation which only approximates what the performance would be in an independent population, with split-sample approaches being the most unreliable and bootstrapping being the most reliable.^44^

As found in an earlier review of hypertension risk prediction models, the FHR model and the Johns Hopkins risk model were still the only models externally validated by authors other than those who developed the models.^5^ There was significant heterogeneity in the FHR model’s discrimination even when stratified by region, suggesting that a model developed in one population does not always perform well in other populations (Supplement 3).

### Risk of bias

In the PROBAST assessment, all models were at high ROB, mainly due to limitations with their analyses (Domain 4). Firstly, most studies either excluded observations with missing data from the analysis (complete-case analysis) or did not report how they handled their missing data. For studies that did not mention missing data at all, it is likely that complete-case analysis was performed because statistical packages exclude observations with missing values unless prompted otherwise. Additionally, complete-case analysis is known to be the most common way of handling missing data in prediction model studies.^9^ We expect that this is due to the researchers not having training in imputation methods or not being aware of the consequences of missing data. Excluding enrolled participants with missing data from the analysis can bias model performance as the analyzed individuals are not a representation of the initial study sample.^9^ This exclusion of enrolled participants due to missing data was also reflected in Domain 4.3.

The second limitation in the analysis domain was the lack of appropriate performance measures (Domain 4.7). Most models in this review reported discriminative performance (AUC) but either did not report any calibration measures whatsoever (n = 19), or only reported statistical tests of calibration rather than plots and tables (n = 17). Among model validation studies, all reported discrimination metrics, and most reported some form of calibration (n = 10). However, only four shared a calibration plot or table. A calibration plot shows the magnitude and direction of miscalibration across the full range of predicted probability, whereas a test like the Hosmer-Lemeshow goodness-of-fit test only presents one overall metric and is very sensitive to sample size and the number of groups. Therefore, the test gives significant results when the sample size is large and nonsignificant results when the sample size is small. Studies that only reported the Hosmer-Lemeshow test received “N” in Domain 4.7. Reporting the AUC without also sharing the calibration gives us an over-optimistic outlook on a model’s performance. Since most participants in a study do not have hypertension, a model with high specificity will result in a high AUC.

The high overall ROB in all studies is cause for concern as study estimates, and therefore the model’s predictive ability, can be flawed. Only one other systematic review of hypertension risk prediction models conducted a bias assessment based on PROBAST and had contrasting results to this review.^8^ The authors concluded that most models were at low ROB even though missing data were handled inappropriately and adequate calibration metrics were lacking in most models (Supplement 4).

### Quality of reporting

Many of the limitations mentioned above and those assessed through PROBAST are not uncommon in prediction model studies. Systematic reviews of prediction models have shown that the quality of prediction model development and validation papers across various outcomes is lacking.^10^ This has led to the development of the TRIPOD statement which was published in 2015.^10^ TRIPOD gives recommendations on the reporting of development and validation of prediction models across all medical domains and types of predictors in order to improve the quality of published prediction model studies. In this review, 19 of 42 models developed, and 2 out of 11 model validations were published after 2015 and could have avoided the issues mentioned above if they had followed TRIPOD recommendations. However, only one model development study met the guidelines.^38^

### Implications

Clinicians and researchers can use models included in this review to identify individuals at high risk of hypertension and subsequently tailor prevention strategies or recruitment in clinical trials. We recommend that users compare models that were developed using data from populations similar to their target population or were shown to have good performance when validated in their target population. For this, it is important to consider the geography, ethnicity, and selection criteria of the model and ensure that it matches the population of interest. For example, a model that excluded women from their sample would not be appropriate to predict hypertension in female patients.

### Strengths and limitations of this review

This review provides a comprehensive summary of studies developing and validating existing risk models for predicting hypertension or increased BP. We used established methodological guidelines for systematic reviews of prognostic models to conduct and report this review.^9,11,45^ We also used PROBAST to assess the ROB and comment on the overall quality of hypertension prediction models. The main limitation of this study is that it included English language studies only. Therefore, we could not compare models published in any non-English studies to those in this review.

## CONCLUSIONS

In this systematic review, we identified and assessed 42 existing models predicting hypertension or increase in BP, as well as 11 external validations of existing models. These models are heterogeneous with respect to predictors included, statistical methods used, definition of predictors and outcome, study population, study design, and sample size. All models have high ROB largely due to limitations in analyses. Existing prediction models should be externally validated in different populations and settings than the ones from the original model. New studies developing prediction models or validating existing models should aim for higher quality by reducing their ROB and using standard reporting guidelines published for risk prediction models.

**Table 5.** Quality assessment of risk of bias and applicability of hypertension prediction models

| Model | ROB |  |  |  | Applicability |  |  | Overall |  |
| --- | --- | --- | --- | --- | --- | --- | --- | --- | --- |
|  | Participants | Predictors | Outcome | Analysis | Participants | Predictors | Outcome | ROB | Applicability |
| 1 | + | + | - | - | + | + | - | - | - |
| 2 | + | + | + | - | + | + | + | - | + |
| 3 | + | + | + | - | + | + | + | - | + |
| 4 | - | - | - | - | + | + | + | - | + |
| 5 | - | - | - | - | + | + | + | - | + |
| 6 | - | - | - | - | + | + | + | - | + |
| 7 | + | + | + | - | + | + | + | - | + |
| 8 | + | + | + | - | + | + | + | - | + |
| 9 | + | + | + | - | + | + | + | - | + |
| 10 | + | + | + | - | + | + | + | - | + |
| 11 | + | + | + | - | + | + | + | - | + |
| 12 | + | + | + | - | + | + | + | - | + |
| 13 | + | + | + | - | + | + | + | - | + |
| 14 | ? | + | - | - | + | + | + | - | + |
| 15 | ? | + | - | - | + | + | + | - | + |
| 16 | ? | + | + | - | + | + | + | - | + |
| 17 | ? | + | + | - | + | + | + | - | + |
| 18 | + | + | + | - | + | + | + | - | + |
| 19 | + | + | + | - | + | + | + | - | + |
| 20 | + | + | + | - | + | + | + | - | + |
| 21 | - | - | - | - | + | + | + | - | + |
| 22 | + | + | + | - | + | + | + | - | + |
| 23 | + | + | + | - | + | + | + | - | + |
| 24 | + | + | + | - | + | + | + | - | + |
| 25 | + | + | + | - | + | + | + | - | + |
| 26 | - | - | - | - | + | + | + | - | + |
| 27 | + | + | + | - | + | + | + | - | + |
| 28 | + | + | + | - | + | + | + | - | + |
| 29 | + | + | + | - | + | + | + | - | + |
| 30 | + | + | + | - | + | + | + | - | + |
| 31 | - | - | - | - | + | + | + | - | + |
| 32 | + | + | + | - | + | + | + | - | + |
| 33 | - | - | - | - | + | + | + | - | + |
| 34 | - | - | - | - | + | + | + | - | + |
| 35 | - | - | - | - | + | + | + | - | + |
| 36 | + | + | + | - | + | + | + | - | + |
| 37 | + | + | + | - | + | + | + | - | + |
| 38 | + | + | + | - | + | + | + | - | + |
| 39 | ? | ? | + | - | + | + | + | - | + |
| 40 | ? | ? | + | - | + | + | + | - | + |
| 41 | ? | ? | + | - | + | + | + | - | + |
| 42 | - | + | + | - | + | + | + | - | + |

ROB = Risk of bias
+ indicates low ROB/low concern regarding applicability
- indicates high ROB/high concern regarding applicability
? indicates unclear ROB/unclear concern regarding applicability

## Data Availability

All data produced in the present work are contained in the manuscript.

## SUPPLEMENT 1: FULL ELECTRONIC SEARCH STRATEGY

### Pubmed

((((“Hypertension” OR “Essential Hypertension”[Mesh] OR Hypertensi*[tiab] OR “blood pressure”[tiab] OR “BP”[tiab] OR prehypertensi*[tiab] OR Pre-hypertensi*[tiab]) AND (“Prediction”[tiab] OR “Predictions”[tiab] OR “Predictive”[tiab] OR “Prognostic”[tiab] OR “Risk assessment” [tiab])) AND (“Multivariate analysis”[tiab] OR “Model”[tiab] OR “Tool”[tiab] OR “Equation”[tiab] OR “Score” OR “Rule”[tiab] OR “rules”[tiab] OR “Regression”[tiab] OR “Machine learning”[tiab] OR “ROC Curve”[tiab] OR “receiver operating characteristic” [tiab] OR “Area under the curve” [tiab])) AND (“Cohort”[tw] OR “observational”[tw] OR “prospective”[tw] OR epidemiolog*[tw])) NOT (“Animals”[mesh] NOT (“Animals”[mesh] AND “Humans”[mesh]))

### Embase

1. (“Hypertension” OR “Essential Hypertension” OR Hypertensi* OR “blood pressure” OR “BP” OR prehypertensi* OR Pre-hypertensi*):ab,ti
2. (“Prediction” OR “Predictions” OR “Predictive” OR “Prognostic” OR “Risk assessment”):ab,ti
3. (“Multivariate analysis” OR “Model” OR “Tool” OR “Equation” OR “Score” OR “Rule” OR “rules” OR “Regression” OR “Machine learning” OR “ROC Curve” OR “receiver operating characteristic” OR “Area under the curve”):ab,ti
4. (“Cohort” OR “observational” OR “prospective” OR epidemiolog*):ab,ti
5. [humans]/lim AND [article]/lim

## SUPPLEMENT 2: FULL LIST OF EXTRACTED ITEMS

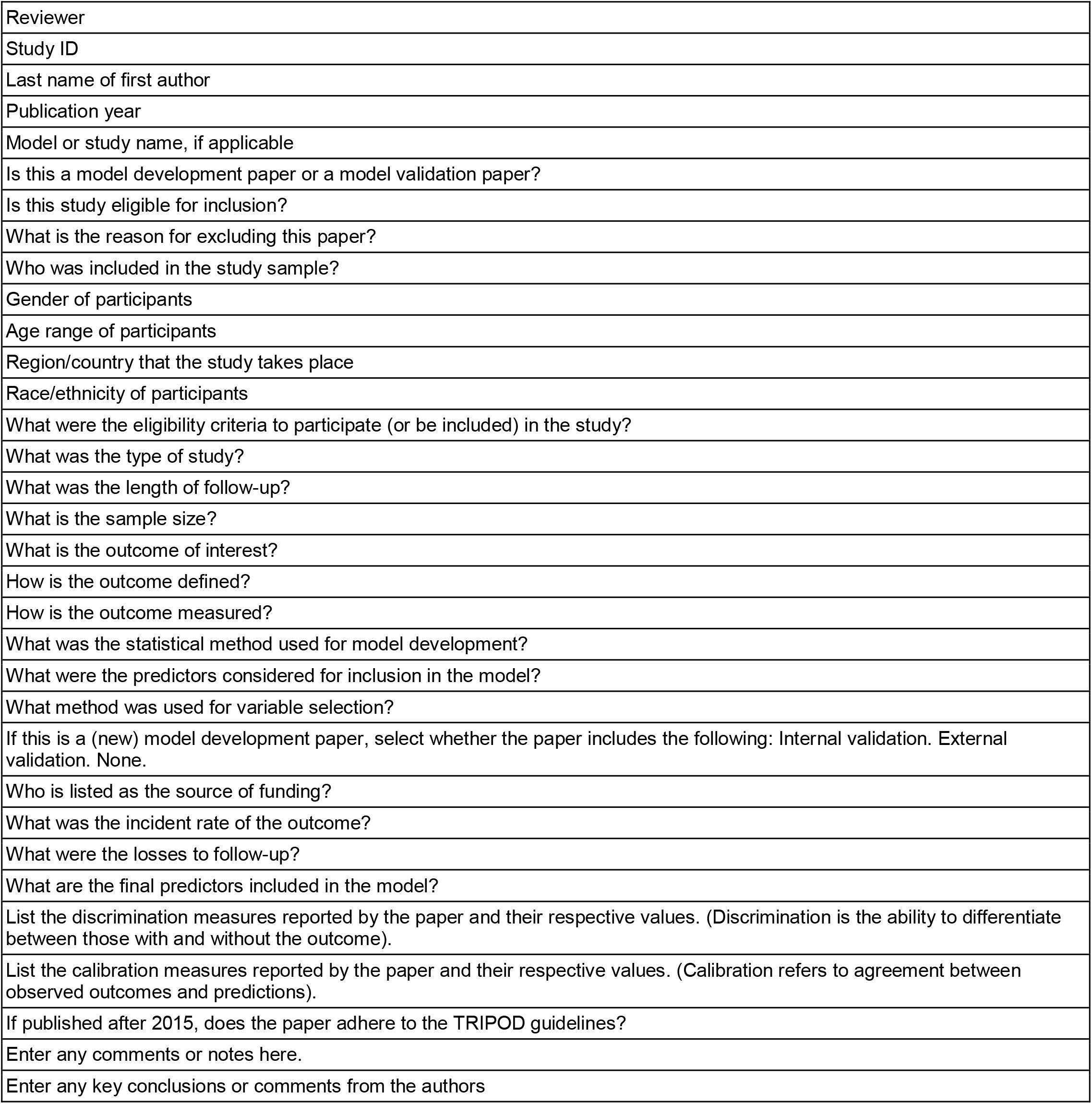

## SUPPLEMENT 3: META ANALYSIS

Forest plot with the AUC for studies that validated the Framingham hypertension risk score, by region.

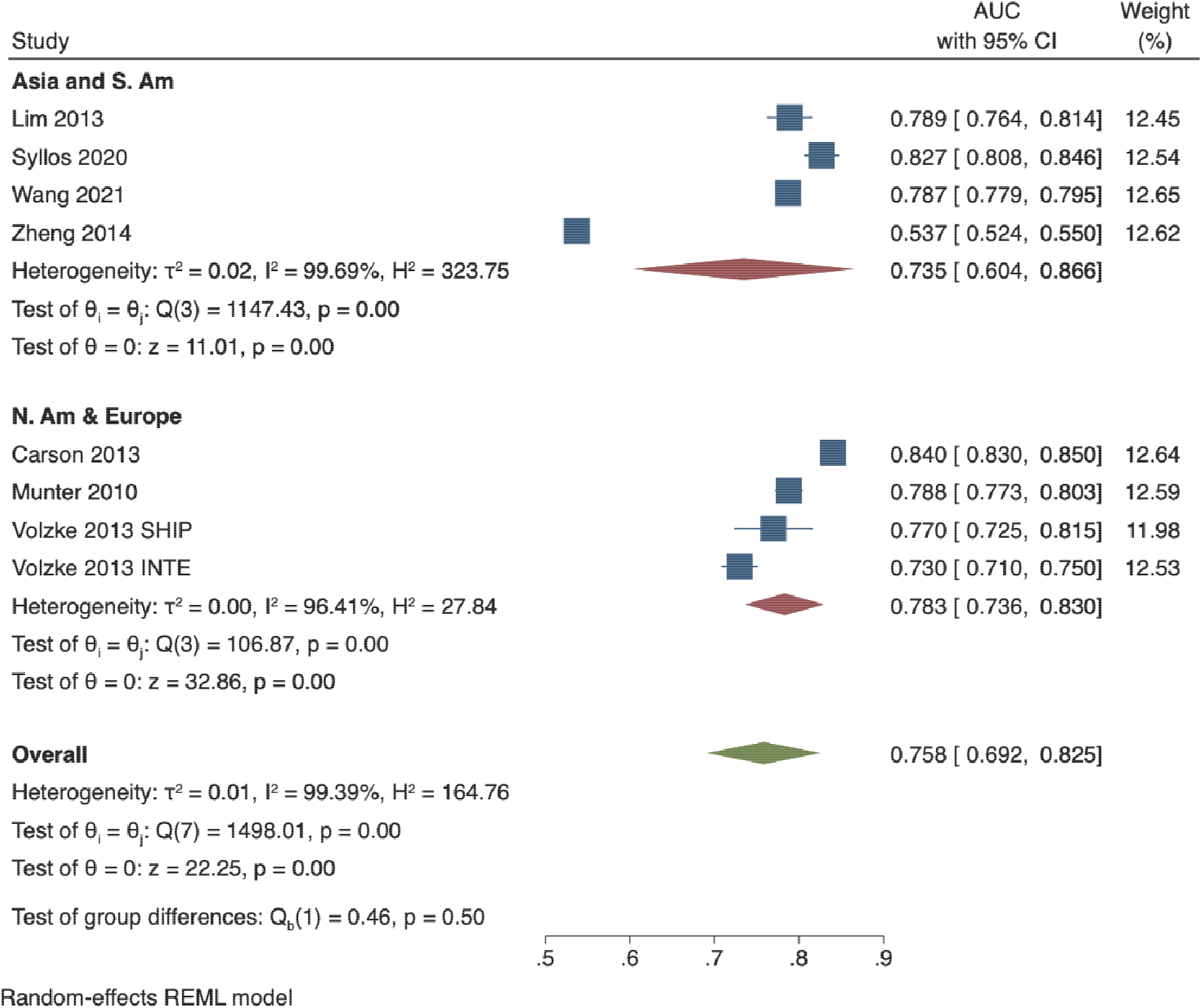

## SUPPLEMENT 4: ROB ASSESSMENT

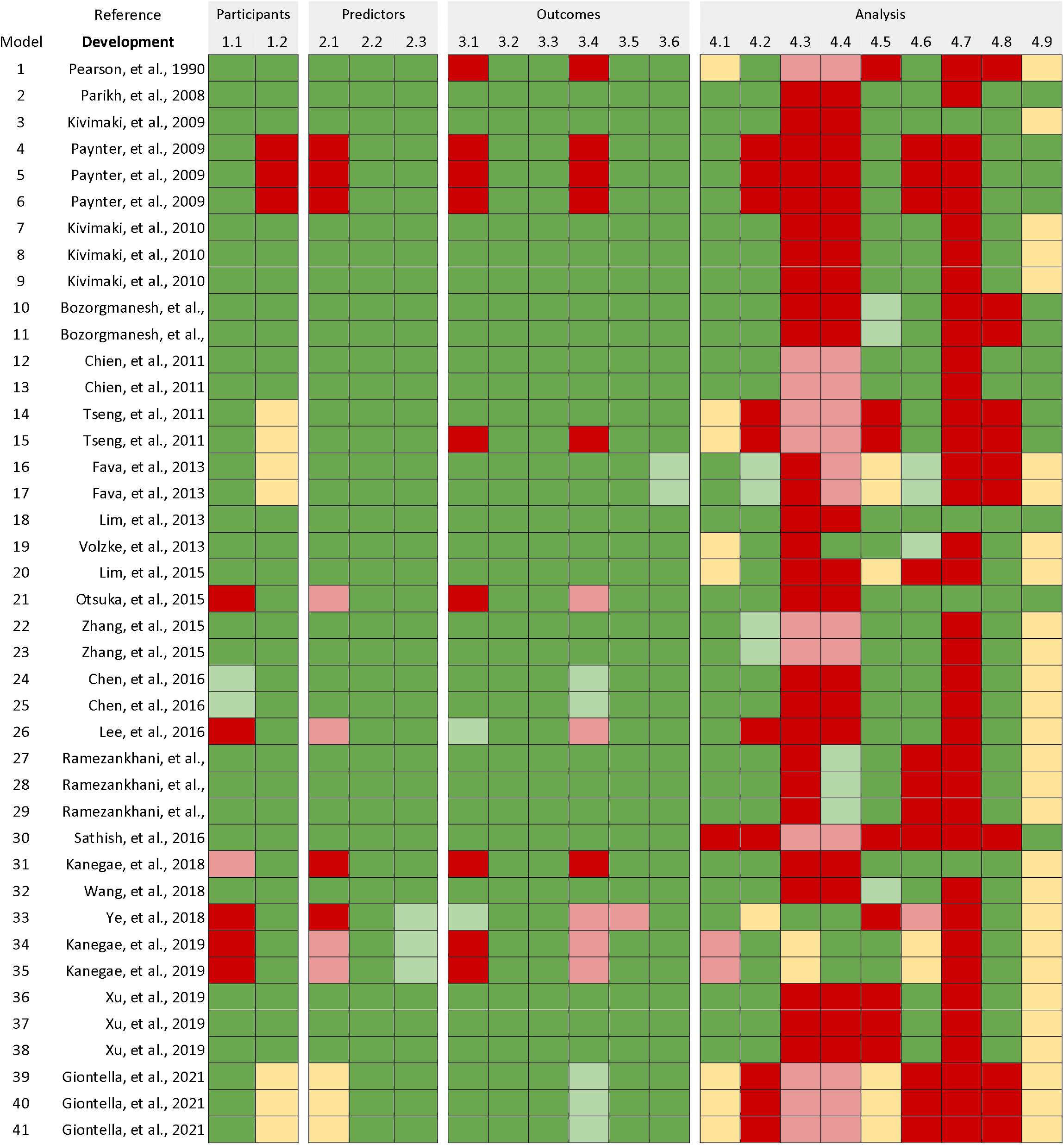
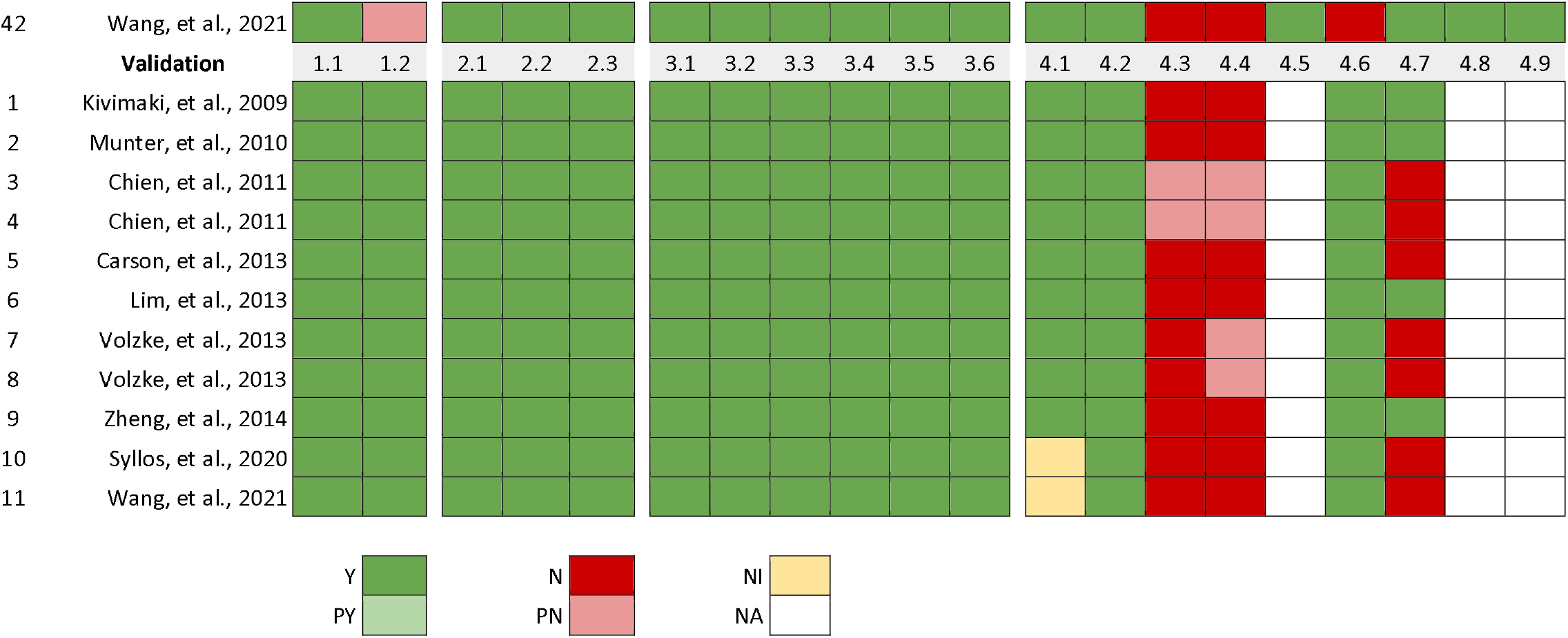

Each signal question was answered as yes (Y), probably yes (PY), no (N), probably no (PN), or no information (NI). NA = Not applicable

**Q1.1:** Were appropriate data sources used, e.g., cohort, RCT, or nested case–control study data?

**Q1.2:** Were all inclusions and exclusions of participants appropriate?

**Q2.1:** Were predictors defined and assessed in a similar way for all participants?

**Q2.2:** Were predictor assessments made without knowledge of outcome data?

**Q2.3:** Are all predictors available at the time the model is intended to be used?

**Q3.1:** Was the outcome determined appropriately?

**Q3.2:** Was a prespecified or standard outcome definition used?

**Q3.3:** Were predictors excluded from the outcome definition?

**Q3.4:** Was the outcome defined and determined in a similar way for all participants?

**Q3.5:** Was the outcome determined without knowledge of predictor information?

**Q3.6:** Was the time interval between predictor assessment and outcome determination appropriate?

**Q4.1:** Were there a reasonable number of participants with the outcome?

**Q4.2:** Were continuous and categorical predictors handled appropriately?

**Q4.3:** Were all enrolled participants included in the analysis?

**Q4.4:** Were participants with missing data handled appropriately?

**Q4.5:** Was selection of predictors based on univariable analysis avoided? [development studies only]

**Q4.6:** Were complexities in the data (e.g., censoring, competing risks, sampling of control participants) accounted for appropriately?

**Q4.7:** Were relevant model performance measures evaluated appropriately?

**Q4.8:** Were model overfitting and optimism in model performance accounted for? [development studies only]

**Q4.9:** Do predictors and their assigned weights in the final model correspond to the results from the reported multivariable analysis? [development studies only]

